# Increased gametocyte production and mosquito infectivity in chronic versus incident *Plasmodium falciparum* infections

**DOI:** 10.1101/2020.04.08.20057927

**Authors:** Aissata Barry, John Bradley, Will Stone, Moussa W. Guelbeogo, Kjerstin Lanke, Alphonse Ouedraogo, Issiaka Soulama, Issa Nébié Ouedraogo, Samuel S. Serme, Lynn Grignard, Katie Patterson, Shehu S. Awandu, Mireille Ouedraogo, Casimire W. Tarama, Désiré Kargougou, Zongo Zoumanaba, Sodiomon B. Sirima, Matthias Marti, Chris Drakeley, Alfred B. Tiono, Teun Bousema

## Abstract

We longitudinally assessed *P. falciparum* parasite kinetics, gametocyte production and infectivity in incident infections that were naturally acquired following infection clearance and in chronic asymptomatic infections in Burkina Faso. 92% (44/48) of the incident cohort developed symptoms and were treated within 35 days, compared to 23% (14/60) of the chronic cohort. All but two individuals with chronic infection were gametocytaemic at enrollment, whereas only 35% (17/48) in the incident cohort developed gametocytes within 35 days. The relative abundance of *ap2-g* transcripts was positively associated with conversion to gametocyte production (i.e. the ratio of gametocytes at day 14 to ring stage parasites at baseline) and was higher in chronic infections. Parasite multiplication rate, assessed by daily molecular parasite quantification, was positively associated with prospective gametocyte production. Most incident infections were cleared before gametocyte density was sufficiently high to infect mosquitoes. In contrast, chronic, asymptomatic infections represented a significant source of mosquito infections. If present, gametocytes were significantly less infectious if concurrent with malaria symptoms. Our observations support the notion that malaria transmission reduction may be expediated by enhanced case management, involving both symptom-screening and infection detection.

## Introduction

The epidemiology of *P. falciparum* transmission stages, gametocytes, is poorly understood. Molecular diagnostics show that low density gametocyte carriage is highly prevalent in endemic populations^1^ and that many low density infections are infectious to mosquitoes^2^, explaining observations from the 1950s that gametocyte free individuals (as determined by microscopy) frequently infected mosquitoes^3^. Few assessments of the population infectious reservoir of malaria parasites have been undertaken^4,5^, particularly with tools capable of detecting low parasite and gametocyte densities^6,7^. Fewer still have been able to assess the association of infectivity with factors other than parasite density, despite clear evidence that gametocyte maturity^8,9^, intrinsic parasite factors^10,11^, human genetic factors^12^, and human clinical and immune responses^13-15^ can have significant influence on the effectiveness of parasite transmission.

The association between malaria symptoms and gametocyte dynamics is particularly poorly understood. Among similarly immune individuals, higher densities of pathogenic, asexual stage malaria parasites are commonly associated with the presentation of symptoms^16^. Because gametocytes develop from asexual parasites, individuals with clinical malaria and higher parasite densities may have more gametocytes and be more likely to infect mosquitoes^17,18^. On the other hand, the slower development of gametocytes may result in higher densities later in infections. This would result in higher gametocyte densities in infections that remain asymptomatic^6,19^. These uncertainties have added to the ongoing debate over the importance of asymptomatic infections for malaria transmission^19-21^. To truly understand the interaction of symptomatic status and the temporal dynamics of malaria infection, longitudinal assessments are required that can determine whether infected individuals living in malaria endemic areas are infectious to mosquitoes before symptoms arise or before malaria infections are detectable by microscopy. Such longitudinal studies can also prospectively assess how gametocyte production is influenced by infection characteristics such as parasite multiplication rates^22^, clinical symptoms, duration of infection and multiplicity of infection^23^.

In the present longitudinal study, we aimed to describe changes in asexual and sexual parasite density and infectiousness at two moments of the parasite’s natural history: immediately after blood stage infection establishment, and during the chronic phase of the infection. We recruited two cohorts of school children from Balonghin, Burkina Faso. In the first cohort, aiming to characterize incident infection dynamics, individuals were cleared of existing infection and monitored weekly by PCR for up to 6 months to detect new infections at their onset. In the second cohort, aiming to characterize chronic asymptomatic infections, individuals with no clinical disease were enrolled for monthly monitoring, and were classified as having chronic and asymptomatic infection when parasites were detected on consecutive visits by PCR in the absence of symptoms. Blood samples were taken to measure total parasite density, multiplicity of infection, parasite multiplication rates over 48h intervals, anti-malarial antibodies, male and female gametocyte densities, and relative *ap2-g* abundance (as a proxy for the sexual conversion rate). Infectivity to mosquitoes was determined upon detection of incident or chronic asymptomatic infections, and at multiple time points during follow-up.

## Results

A total of 253 individuals were screened for participation in the incident infection sub-study; 80 were confirmed *Plasmodium* negative by nested PCR (nPCR), willing to participate, and met all other criteria for enrolment (**table 1**). Incident infections were detected by nPCR for 65% (52/80) of individuals after a median participation of 27 days (interquartile range [IQR] 20-41). After retrospective qPCR assays were performed on all pre- and post-enrolment samples, 4 individuals were excluded from subsequent analysis because they had parasites measured by qPCR more than 2 weeks before they were detected by nPCR, leaving 48 individuals in the incident infection cohort.

**Table 1:** Characteristics and parasite metrics of participants

| Cohort | Year | N | Female, n (%) | Age, median (range) | Asexual density at recruitment, median (IQR) | Peak asexual density, median (IQR) | Symptoms before day 35, n (%) | Number of clones, median (IQR) | Gametocyte positive at recruitment, n (%) | Gametocyte positive before day 35, n (%) | Peak gametocyte density (positives), median (IQR) |
| --- | --- | --- | --- | --- | --- | --- | --- | --- | --- | --- | --- |
| Incident | 2015 | 31 | 10 (32%) | 7 (5,10) | 85.7, (1.6-432.5) | 10839 (2529-71116) | 30 (97%) | 3 (1-5) | 1 (3%) | 9 (29%) | 0.08 (0.05-0.16) |
|  | 2017 | 17 | 8 (47%) | 8 (5,10) | 201.9 (28.1-587.6) | 14193 (5689-21089) | 14 (82%) | 2 (2-4) | 2 (12%) | 8 (47%) | 0.32 (0.05-41.3) |
|  | Total | 48 | 18 (38%) | 7 (5,10) | 127.1 (2.7-561.7) | 11593 (3373-40345) | 44 (92%) | 3 (1-5) | 3 (6%) | 17 (35%) | 0.12 (0.05-0.84) |
| Chronic | 2016 | 40 | 17 (43%) | 8 (6,10) | 337.6 (91.2-1300.4) | 2320 (1067-8678) | 10 (25%) | 6 (3-8) | 38 (95%) | 40 (100%) | 15.3 (6.5-26.7) |
|  | 2017 | 20 | 9 (45%) | 9 (6,10) | 172.6 (84.2-386.4) | 1421 (607-3644) | 4 (20%) | 7 (5-8) | 20 (100%) | 20 (100%) | 7.8 (2.6-41.1) |
|  | Total | 60 | 22 (42%) | 8 (6,10) | 295.2 (92.2-796.3) | 1987 (770-5599) | 14 (23%) | 7 (4-8) | 58 (97%) | 60 (100%) | 13.4 (5.2-28.2) |

For the chronic infection sub-study, 228 individuals were screened and 60 were enrolled, having been confirmed as asymptomatic, and parasite positive for at least 1 month prior (median 28.5 days [IQR 28-38.5]). Individuals with confirmed incident or chronic infections were followed daily for 1 week and weekly until day 35 or the presentation of symptoms (**figure 1**). Genotyping of merozoite surface protein-2 (MSP2) demonstrated that the majority of cohort participants had clonally complex infections at the start of intensive follow-up, both for incident infections (74% [29/39] multi-clonal) and chronic infections (100% [57/57] multi-clonal) (**table 1**).

**Figure 1:**
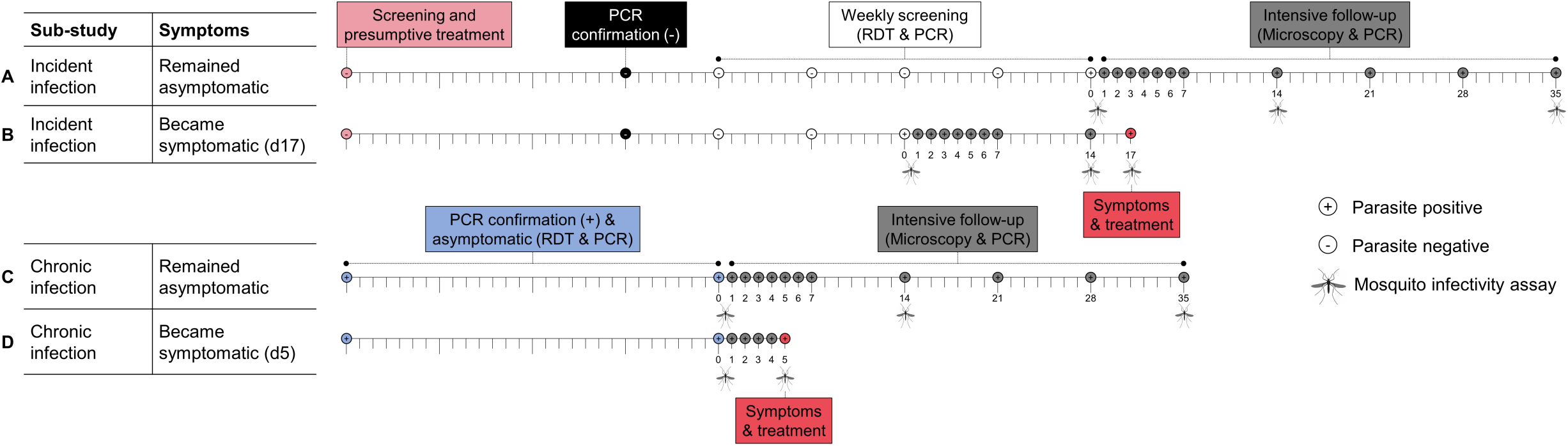
Illustrative follow up patterns. Four illustrative follow up patterns are shown; two for each of the sub-studies. Tick marks represent days and weeks, and sampling/screening time points are shown as circles. + and – inside circles indicates requirements for parasite status, determined as indicated. In the incident infection sub-study (**A** and **B**), individuals were screened to confirm parasite negativity by microscopy, presumptively treated for sub-patent infection (light pink circle), and screened 3 weeks later with qualitative nested PCR based on detection of *Plasmodium* specific 18s ribosomal DNA (nPCR) (black circle) to confirm the absence of sub-patent infection. They were then screened weekly with nPCR (white circles) until the onset of an infection. Intensive follow up (grey circles) proceeded every day for 1 week, and weekly until day 35 after parasite detection. Study participants were closely monitored for the development of malaria symptoms. Artemether-lumefantrine anti-malarial treatment was given upon the detection of symptoms or 35 days after initial detection of infection by nPCR, whichever came first. Individuals in the chronic infection cohort were screened monthly with nPCR to confirm chronic infection (blue circles), defined as two sequential visits with confirmed parasitaemia by nPCR without any symptoms of malaria disease. Intensive follow-up proceeding from confirmation of chronic infection (grey circles) was as for incident infections. Mosquito feeding was conducted in both sub-studies at the onset of intensive sampling (day 0), and at day 14 and 35 in the absence of symptoms. Mosquito feeding assays were conducted on the day of symptom detection if applicable.

### Infection dynamics

Total parasite and gametocyte density trajectories for all individuals are presented in **supplemental figure 2**, with values relative to the time of first detection shown in **figures 2A** and **2B**. The majority of individuals with incident infections experienced symptoms prior to day 35 (92%, 44/48), compared to only 23% (14/60) of those with chronic infections. Median peak total parasite density (throughout follow-up) was higher in incident infections (p=0.0001) although when participants entered the intensive follow-up for either cohort, total parasite densities were similar (**table 1**). For those who developed symptoms, the median interval between the start of the intensive follow-up and the onset of symptoms was 4 days (IQR 2 - 7) for incident infections, and 11 days (IQR 6-22) for chronic infections. In chronic infections that became symptomatic, total parasite densities increased during follow-up (average increase 6.0% per day [CI 0.08 – 15.2, p=0.023, n=14]), whereas for those that remained asymptomatic total parasite density decreased (average decrease 1.7% per day [CI 0.6 - 2.7%, p=0.002, n=46; p value for difference <0.0001]). Gametocyte densities decreased by on average 1.9% per day in chronic infections [CI 1.1 – 2.8, p<0.0001, n = 60] with the rate of decrease similar among infections that became symptomatic and those that remained asymptomatic (p=0.067).

**Figure 2:**
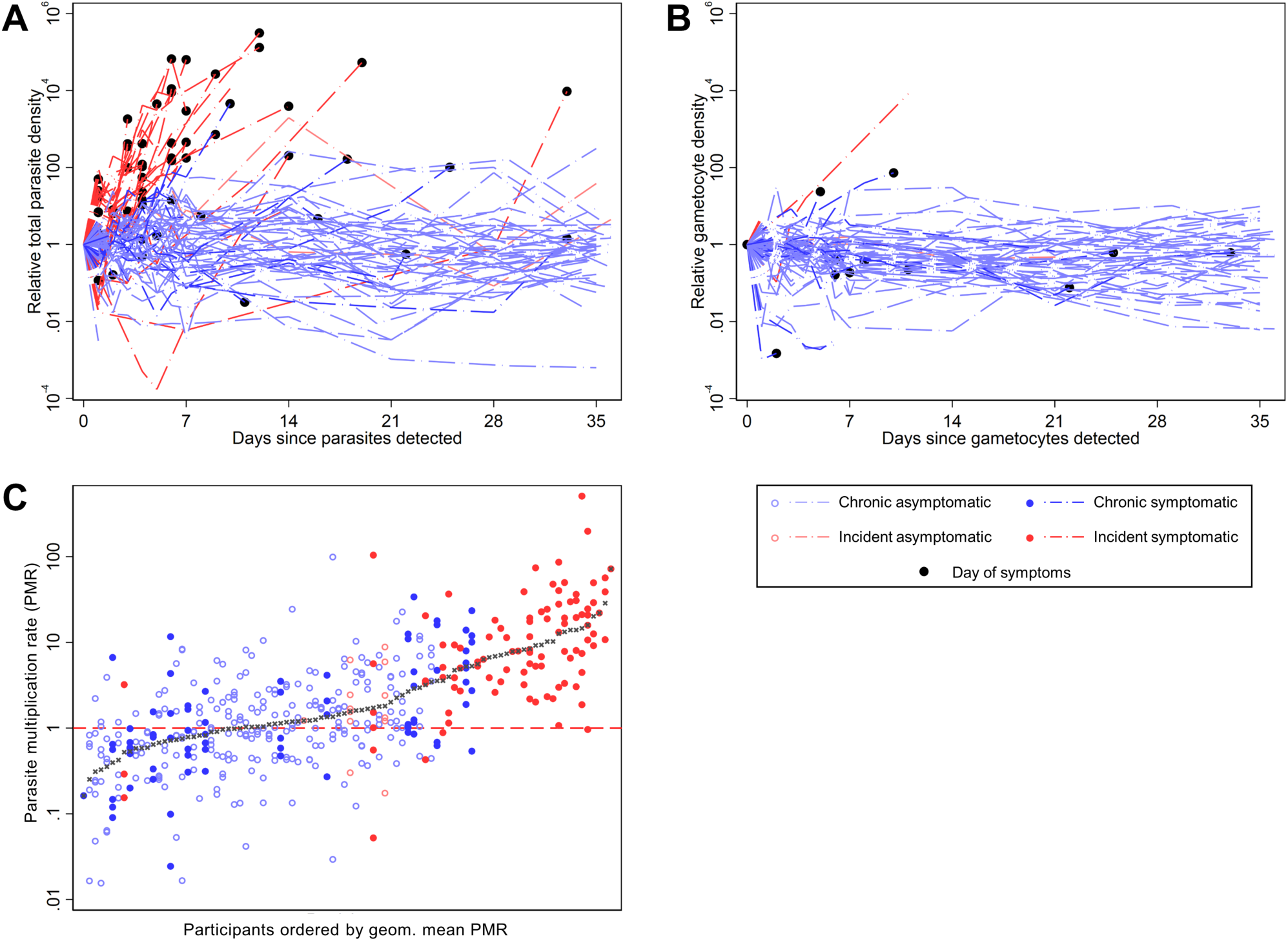
Total parasite and gametocyte dynamics. In **A** and **B** total parasite and gametocyte densities are presented as values relative to the time point of their first detection (y = the number of days since first detection). In **C**, total parasite multiplication rates (PMR) were calculated as the change in density between observations over 48-hour intervals in the first 7 days of intensive follow up. Participants are ordered by the geometric mean of their MR observations. 431 measurements of total parasite MR from 92 participants where available (median per person = 6 [IQR 3,6]). Incident asymptomatic: 13 observations (3 individuals), median observations = 6, IQR (1,6); Acute symptomatic: 98 observations (31 individuals), median observations = 3, IQR (2,4); Chronic asymptomatic: 247 observations (44 individuals), median observations = 6, IQR (6,6); Chronic symptomatic: 73 observations (14 individuals), median observations = 6, IQR (5,6).

The total parasite multiplication rate (PMR) during 48-hour intervals (i.e. parasite density at 48 hrs/parasite density at baseline) in the first week of intensive follow up was estimated for each available interval for each individual (**figure 2C**). Geometric mean PMR was higher in individuals with incident infection (6.70, 95% CI [4.76-9.42]) compared to those with chronic infection (1.08 [0.90, 1.28]) (p < 0.0001); age and sex had no significant effect on this association. PMR was higher in incident infections leading to symptoms than incident infections that remained asymptomatic (p=0.001) or chronic infections with (p<0.001) or without symptoms (p<0.001). There was no evidence that PMR was different between incident infections that remained asymptomatic, chronic infections that remained asymptomatic, and chronic infections that became symptomatic (p=0.301). Neither multiplicity of infection (p=0.565) nor Hb genotype (p=914) were associated with total parasite MR, and there was no evidence that PMR changed with time during the week of intensive follow-up (p=0.866).

The magnitude of antibody responses specific to 12 *P. falciparum* antigens was determined at the start of the screening for incident infections (i.e. before infection) (n=46), or at the time of entry into the chronic cohort (n=51). Antibody levels were uniformly higher in samples collected from individuals with chronic infections (p<0.0001 for all 12 antigens). Baseline antibody levels were significantly higher in individuals who remained asymptomatic for MSP2-ch150 (p=0.036) in incident infections, and EBA-181 (p=0.042) and GLURP-R2 (p=0.032) in chronic infections (**figure 3, supplemental table 1**). In incident infections antibody levels were negatively associated with PMR for EBA-140 (p<0.001), EBA-181 (p=0.037), and PfAMA-1 (p=0.030).

**Figure 3:**
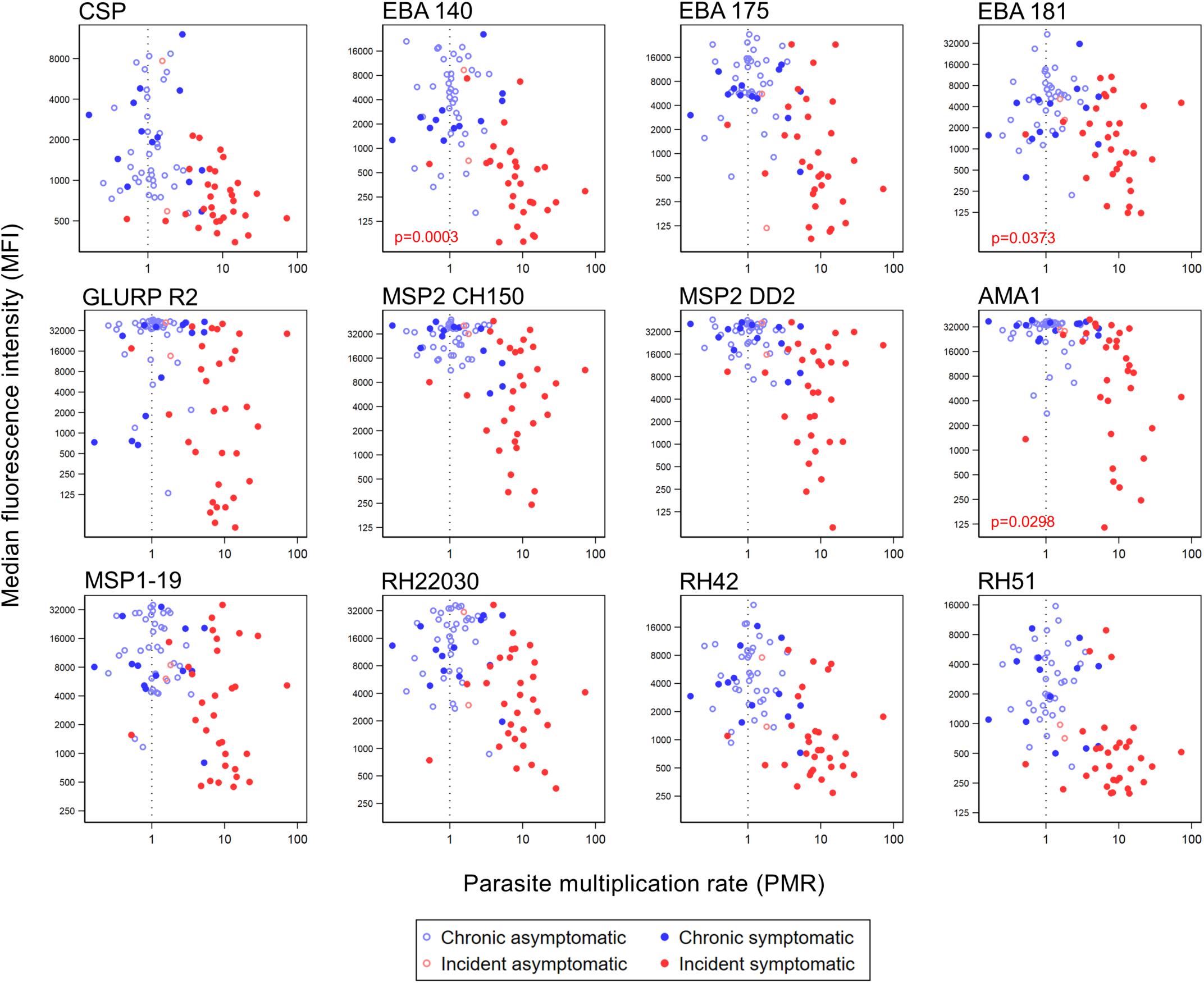
Magnitude of antibody response and parasite multiplication rate. Magnitude of antibody response to specific *P. falciparum* antigens is given as the background adjusted median fluorescence intensity (MFI). Parasite multiplication rate (PMR) is presented as the geometric mean of each individuals PMR observations, with a dashed line at PMR=1 (no change). Samples were assayed from the start of the screening for incident infections (i.e. before infection) (n=46), or at the point of confirmation of chronic infections (n=51).

Among individuals with chronic infections, antibody levels had no apparent association with mean PMR.

### Gametocytes and infectivity

Gametocytes were detected by qRT-PCR in 6% (3/48) of individuals with incident infection at first detection of infection, and in 97% (58/60) of individuals with a qPCR confirmed chronic infection. Only 35% (17/48) of individuals in the incident cohort developed detectable gametocytaemia during their follow-up. Median peak gametocyte density was significantly higher in chronic infections (p<0.0001). Whilst there was considerable variation between individuals in gametocyte densities (**figure 4A**), densities within individuals were relatively stable and the standard deviation of the gametocyte density change over 48 hr intervals was significantly lower than PMR in the same individuals (p<0.0001) (**supplemental figure 3A**); among chronic infections, gametocyte densities slightly decreased in the first week of observation (mean change over 48hr intervals = 0.87 [95% CI 0.77-0.97]).

**Figure 4:**
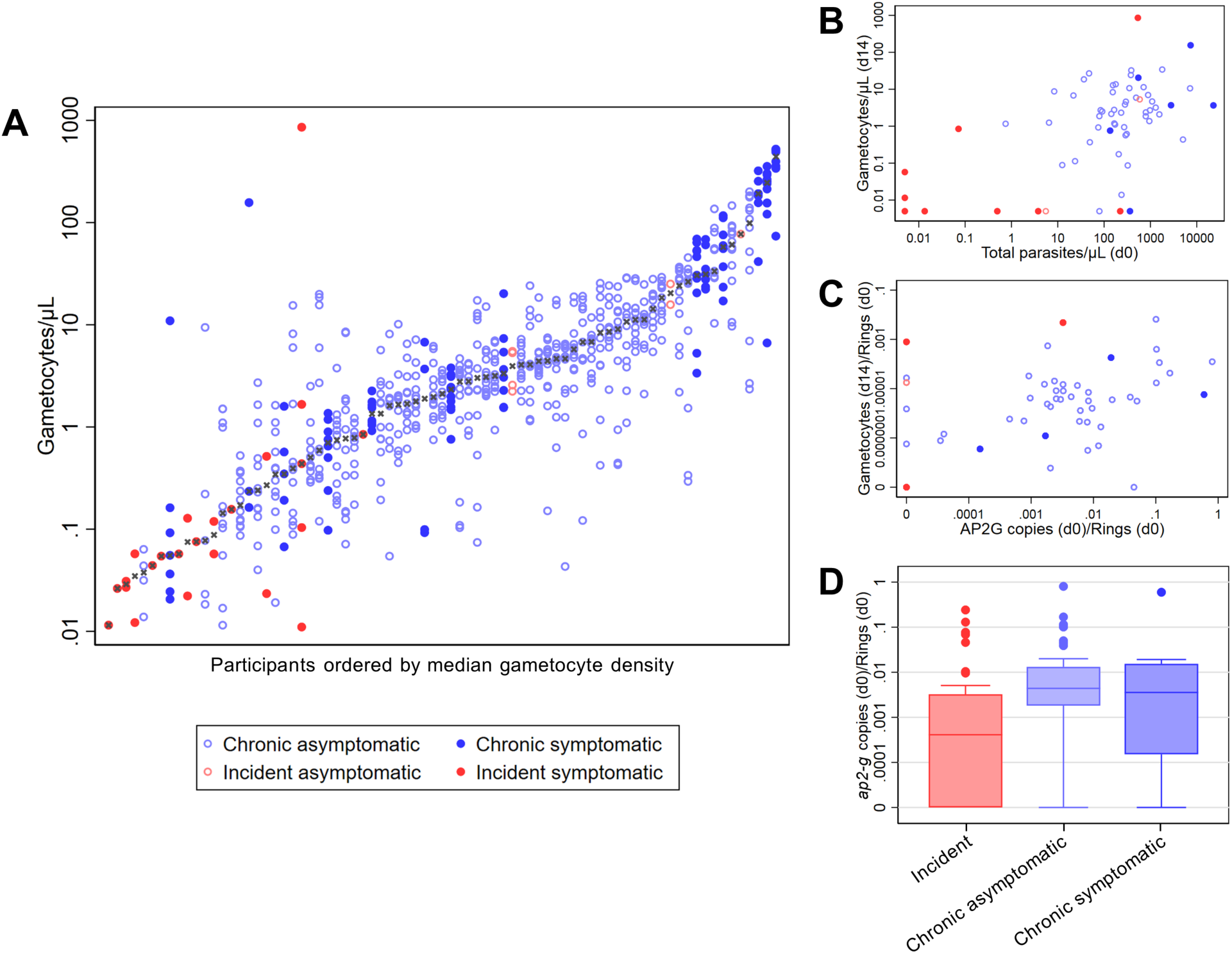
Gametocytes and gametocytogenesis. Timepoints are given as the number of days after enrolment into the incident or chronic infection cohorts. **A**. Gametocyte density measures from all gametocyte positive individuals (positive at any time point during follow-up). There were 928 measurements of gametocyte density in total [median per person =4, IQR (2-8)]. Individuals are ranked by median gametocyte density. **B**. The association of gametocyte density at day 14 and total parasite density at enrolment (day 0). **C**. Gametocyte fraction and sexual commitment. Gametocyte fraction on the y-axis is given as the ratio of gametocytes per µL on day 14 to ring stage parasites per µL on day 0. Sexual commitment was measured as the ratio of ap2-g mRNA copies to the density of ring stage parasites/ µL at day 0. **D**. Sexual commitment and infection characteristics. Sexual commitment is shown as in B.

Combining both cohorts, the association of gametocyte densities at day 14 was significantly stronger with total parasite densities at day 0 (Spearman correlation 0.48 p=0.0001) (**figure 4B**) than at day 14 (Spearman correlation -0.19 p=0.1292) (**supplemental figure 3B**). PMR and total density at enrolment were independently positively associated with day 14 gametocyte density whilst gametocyte densities were lower among individuals with incident infection and among older individuals (**table 2**).

**Table 2:** Predictors of gametocyte density on day 14. Data on day 14 gametocyte density was available for 65 individuals, 52 with chronic infections and 13 with incident infections. Hb genotype = Haemoglobin genotype. MOI = Multiplicity of infection (number of parasite clones). Aggregate magnitude of antibody response was calculated by ranking each individual according to their combined response to 13 *P. falciparum* antigens, as described in the methods. *Adjusted for the whether the infection was incident. †Adjusted for the whether the infection was incident, log total parasite density/µl on day 0, log total parasite multiplication rate (PMR), and age.

| Covariate | Difference in log gametocytes/μl on day 14* | Adjusted difference in log gametocytes/μl on day 14† |
| --- | --- | --- |
| Incident infection | -4.07 [-5.96, -2.18], p<0.001 | -2.52 [-4.55, -0.50], p=0.016 |
| Female | -0.95 [-2.30, 0.40], p=0.163 | - |
| Age |  |  |
| 5-7 | 0 | 0 |
| 8-10 | -1.75 [-3.30, -0.19], p=0.028 | -2.11 [-3.44, -0.77], p=0.002 |
| Log total parasite density/μl on day 0 (1 unit increase) | 0.38 [0.13, 0.63], p=0.004 | 0.52 [0.27, 0.77], p<0.001 |
| Log total parasite density/μl on day 14 (1 unit increase) | 0.10 [-0.14, 0.35], p=0.411 | - |
| Log PMR (1 unit increase) | 1.06 [0.05, 2.07], p=0.040 | 1.57 [0.08, 3.07], p=0.040 |
| Hb g/dl on day 14 (1 unit increase) | 0.29 [-0.22, 0.81], p=0.256 |  |
| Hb genotype | p=0.931 |  |
| AA | 0 | - |
| C | 0.31 [-1.36, 1.97] | - |
| S | 0.67 [-1.40, 2.73] | - |
| SC | 0.52 [-4.85, 5.89] | - |
| MOI | p=0.908 |  |
| 1-3 | 0 | - |
| 4-6 | 0.16 [-1.97, 2.30] | - |
| 7-14 | 0.41 [-1.61, 2.44] | - |
| Aggregate magnitude of antibody response | p=0.285 |  |
| Lowest tertile | 0 | - |
| Middle tertile | 0.40 [-1.33, 2.14] | - |
| Highest tertile | 2.27 [-0.57, 5.11] | - |

The ratio of gametocytes/µL on day 14 to rings/µL on day 0 (representing the proportion of asexual parasites at baseline that converted to gametocytes during this period) was strongly associated with the relative abundance of *ap2-g* on day 0 (as a proxy for the sexual conversion rate) (Spearman correlation coefficient 0.42, p=0.0015) (**figure 4C**). Relative *ap2-g* abundance on day 0 was significantly higher in chronic infections than in incident infections (p<0.0001) (**figure 4D & supplemental figure 4**).

212 mosquito feeding assays were conducted with 52 assays in the incident cohort and 160 assays in the chronic cohort (median of 79 [IQR 71-82] mosquitoes dissected per assay). Overall, there was a positive association between gametocyte density and mosquito infection rate (**figure 5A**, Spearman correlation 0.51, p<0.0001)^24^. In assays on participants with incident infections 0.1% (5/3,403) of mosquitoes became infected, compared to 4.5% (554/12,405) in assays conducted on individuals with chronic infection. Gametocyte densities were generally higher in chronic infections, but after adjustment for gametocyte density the risk of mosquito infection from individuals in the chronic cohort remained higher (risk ratio = 10.44 [95% CI 2.79-39.01], p = 0.0005) (figure 4A). After adjusting for gametocyte density and cohort, the risk of infecting a mosquito at any point was not lower in infections that became symptomatic. However, when feeding assays were conducted concurrently with the presentation of symptoms (25/212) infectivity was signficantly decreased (risk ratio = 0.30 [95% CI 0.13-0.71], p = 0.0061). During chronic infections there was no significant effect of the day (after enrollment) on which feeding assays were performed on infectivity, with (p=0.332) or without (p=0.058) adjustment for gametocyte density (**figure 5B**). There was no significant correlation between total parasite density and infectivity (Spearman correlation 0.12, p=0.081), however, 88% of infectious individuals had total parasite densities above 50/µL (between the detection thresholds of expert field microscopy and standard RDTs ^25^) (**supplemental figure 5**). 100% of infectious individuals had parasites above the estimated detection threshold of standard qualitative PCR (1 parasite/ µL)^26^.

**Figure 5:**
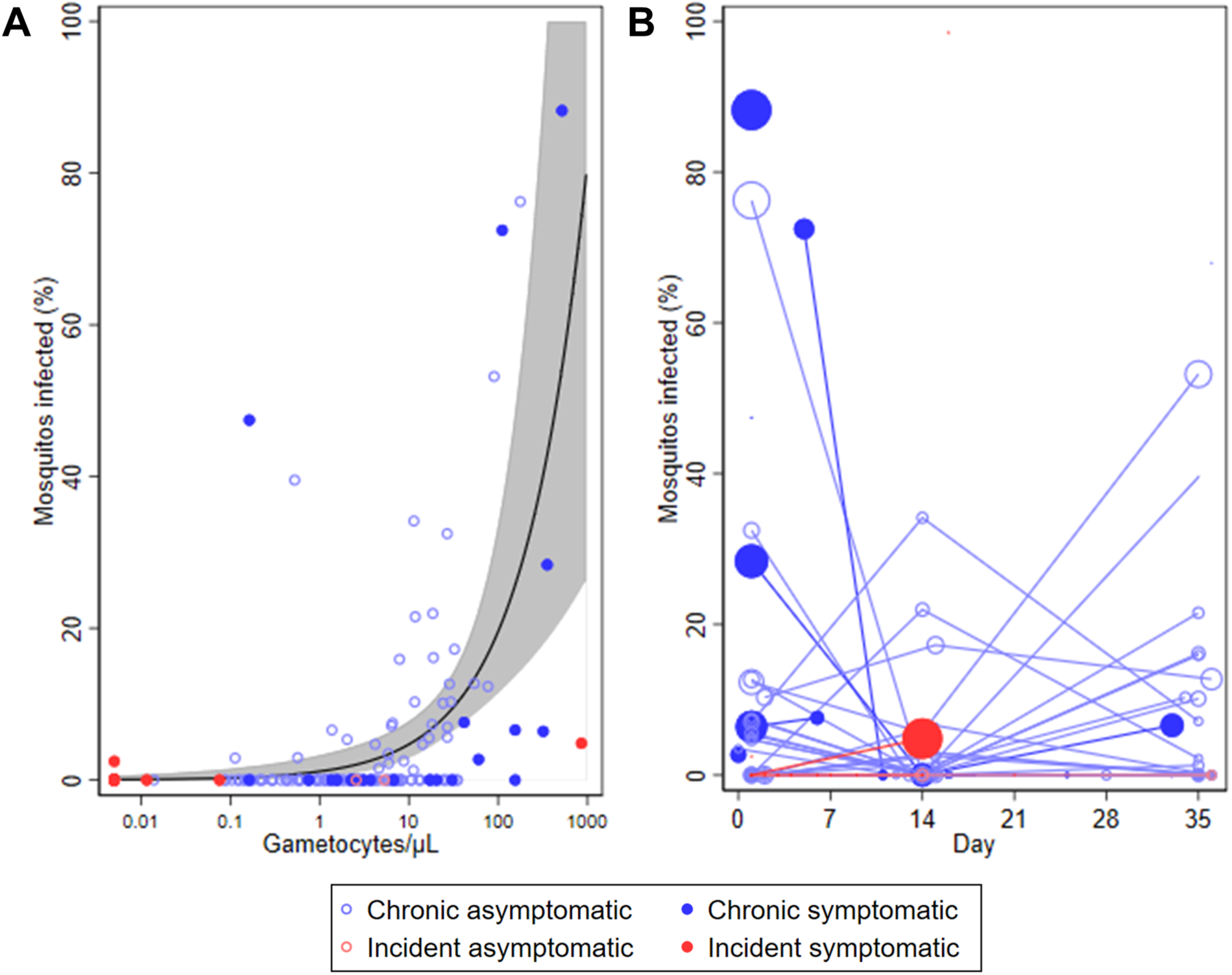
Mosquito infectivity and gametocyte density. **A**. The line indicates the shape of the model for the association of gametocyte density and percent of mosquitoes infected in mosquito feeding assays for individuals with chronic infections. This model is described in detail elsewhere^24^. Grey shading represents the 95% confidence interval for the model. **B**. The percentage of mosquitoes infected over time, shown as days since the start of intensive follow up. The size of the data points is proportional to the log gametocyte density in blood samples taken concurrent with the mosquito feeding assay.

Gametocyte sex ratio (analysed as the percentage of total gametocytes that were male) was significantly higher in chronic infections (median 38.1%) than incident infections (20.4%) (p=0.027) (**supplemental figure 6**). Sex ratio did not differ between chronic infections that became symptomatic and those that remained asymptomatic throughout follow up (p=0.606). There was no association between Hb density at enrolment and gametocyte sex ratio at day 14 for participants with incident infections (p=0.482), chronic asymptomatic infections (p=0.620), or chronic infections that became symptomatic (p=0.116).

## Discussion

To maximise the efficacy of malaria control and elimination programs, there is a need to identify which individuals are most important for transmission of *Plasmodium* to mosquitoes. Among our study population of school-aged children with intensively monitored incident and chronic *P. falciparum* infections, we observed marked variation in gametocyte production and mosquito infectivity. Incident infections were characterized by high total parasite multiplication rates and low gametocyte commitment. More than 90% of incident infections resulted in a detectable fever within 35 days, which in most cases resulted in infections with gametocyte densities insufficiently high to infect mosquitoes. Gametocytes appearing early upon infection and when an individual was symptomatic were less infectious to mosquitoes compared to those in chronic infections.

In the current study, children with incident and chronic infections were examined in detail for parasite kinetics and infectivity to mosquitoes. The study was conducted in an area of intense malaria transmission, confirmed by the observation that 65% (52/80) of parasite free individuals became nPCR positive within 4 weeks (median participation 27 days [IQR 20-41]). Incident infections were frequently clonally complex, indicating either co-transmission of multiple *P. falciparum* strains during mosquito bites^27,28^ or repeated inoculations over a short time-period. Though all participants are likely to have experienced multiple clinical malaria episodes prior to enrolment, the rapid infection rates in our study argue against the existence of sterile immune protection^29,30^. However, the high proportion of individuals with chronic infections who remained asymptomatic highlights substantial clinical immunity in this population age-group^31^. Antibody responses to specific blood stage malaria antigens including EBA^32^, AMA-1^33^ and MSP-2^34,35^ were associated with the ability to control parasite multiplication and remain symptom-free. In our incident infection cohort, the majority of individuals failed to control parasite multiplication and developed malaria symptoms (i.e. fever). Importantly, incident infections occurred after ensuring individuals were free of infection by PCR. In Mali, baseline parasitaemia did not influence the risk of developing clinical malaria^36^. It nevertheless remains to be confirmed whether the likelihood of detecting incident infections by repeated fever screening, that was high in the current study, is influenced by clearance of infections before the transmission season. Contrary to our expectations, haemoglobin genotype was not associated with parasite multiplication rates or the likelihood of remaining free of malaria-symptoms upon infection^37,38^. This may have been due to our understandably conservative definition of symptomatic malaria; the protection associated with HbS increases with the severity of disease^39^, while in our study individuals were activley followed and treated immediatley upon presentation of a mild fever or reported fever in the last 24 hours. Our finding that HbS did not affect total parasite MR is in line with a controlled human infection study in Gabon where only the likelihood of symptoms and not MR was lower for HbS individuals compared to those with normal haemoglobin^40^.

Sexual conversion rates, quantified by relative *pfap2-g* abundance^41^, were higher in chronic infections than incident infections. This suggests that although gametocyte production may start as early as during the first wave of erythrocytic shizogony^42,43^, investment in asexual parasite multiplication dominates the early phase of infection while chronic infections are marked by higher investment in gametocyte production. This may be due to chronic inflammation and hence reduced levels of Lysophosphatidylcholine (LysoPC) continuously triggering elevated rates of gametocyte production^44^. Our observation that after adjusting for gametocyte density, the risk of mosquito infection was lower in incident infections suggests that gametocytes arising in the incident cohort were less mature at the time of feeding. The requirement for gametocyte maturation following release into the circulation would be in line with earlier *in vitro* observations that morphologically mature gametocytes may require several days of maturation to reach peak infectivity^45^ and with findings in controlled human infections where infectivity is only observed several days after gametocyte densities plateau^46^.

Importantly, many children in the incident infection cohorts developed symptoms of clinical malaria early upon infection. When symptoms were detected only 5% (2/42) of individuals had detectable gametocytes, with densities of 0.06/µL and 0.85/µL. Among chronic infections, marked variation in gametocyte production was observed that was positively associated with preceding total parasite density and multiplication rates, and negatively associated with age. The lower gametocyte production among older children was not explained by antibody responses to blood-stage antigens, potentially suggesting a role for recently described immune responses that reduce gametocyte maturation and increase with age^47^.

The low gametocyte production among incident infections and the rapid abrogation of infection due to early treatment resulted in marked differences in the likelihood of mosquitoes becoming infected; only 0.1% of mosquitoes became infected in incident malaria infections compared to 4.5% in chronic malaria infections. Whilst this is largely explained by the number of gametocytes present in the blood at the time of feeding, we also observed that mosquito infection rates were lower from gametocytaemic blood samples from individuals with malaria symptoms^48^. This is in line with earlier findings, principally from animal models, on gametocyte-inactivating activity of inflammatory cytokines and reactive intermediates^49,50^.The mechanism by which this might be achieved and the relative importance for malaria transmission epidemiology requires further study.

From a public health perspective, our findings indicate that there is a window of opportunity to prevent onward malaria transmission from incident infections in semi-immune populations. The abundance, poor detectability, and infectiousness of asymptomatic malaria infections suggest that these may be a considerable stumbling block for malaria elimination. However, if most infections are initally symptomatic (i.e. if the chronic infections we observed reflect the tail-end of symptomatic incident infections) enhancing case management to maximize accessibility of diagnosis and care may abrogate infections early on and, potentially, before individuals become infectious^51^. This is additionally supported by our observations that incident infections have initially low gametocyte production, are detectable by rigorous symptom screening, and that gametocytes are less likely to achieve mosquito infection when arising early upon infection or when sampled during a clinical episode. Enhanced case management may thus reduce the proportion of infections that proceed to become highly infectious^52^, in addition to the obvious clinical benefits of preventing progression to severe disease.

## Methods

### Study location and screening

The study protocol was approved by the ethical review boards of the London School of Hygiene and Tropical medicine (LSHTM) (#9008), the Centre National de Recherche et de Formation sur le Paludisme (CNRFP) (Deliberation numbers 2015-3-033) and the Burkina Faso National Ethical Committee for Health Research.

The study was conducted between June 2015 and December 2017 in Balonghin, in the health district of Saponé 45 kilometres Southwest of Ouagadougou, Burkina Faso. Balonghin experiences intense and highly seasonal malaria transmission from June to October. Children aged ≥5-10 years were screened for enrolment in the incident infection cohort in 2015 and 2017, and the chronic infection cohort in 2016 and 2017. 7/108 individuals participated in the study in two years, with the remainder participating in a single year (see *supplemental information*).

The purpose of the study and practical consequences of participation were explained during community meetings; screening was subsequently conducted at local schools. Individuals providing consent (>12 years) or whose parent or guardian provided consent (<12 years) for screening were examined by a GCP-trained clinician. General criteria for enrolment in either cohort were that individuals were aged ≥5-10 years, were willing to provide repeated blood samples over a 6-month period, and if their caregivers provide informed consent. Exclusion criteria were complicated symptomatic malaria (defined according to standard World Health Organization criteria), anaemia (Hb<8g/dL), presence of any (chronic) illness that requires immediate clinical care, family history of sudden death or of congenital or clinical conditions known to prolong QTc interval (e.g. family history of symptomatic cardiac arrhythmias, clinically relevant bradycardia or severe cardiac disease), current treatment with drugs which could induce a lengthening of QT interval, known history of hypersensitivity, allergic or adverse reactions to piperaquine or other aminoquinolones, severe malnutrition (weight-for-height being below -3 standard deviation or less than 70% of median of the NCHS/WHO normalized reference values), weight below 15 kg, and current or previous participation in malaria vaccine trials.

### Sampling: Incident infection cohort

During screening for the incident infection cohort, a blood smear was taken for malaria diagnosis. Individuals were eligible for enrolment if microscopy negative and were treated presumptively with Dihydroartemisinin-Piperaquine (Duocotexin®, Beijing Holley-Cotec Pharmaceutical, China, 40 mg dihydroartemisinin and 320 mg piperaquine tetra-phosphate per tablet) to clear possible sub-patent infections. Following confirmation of malaria parasite negativity 3 weeks later by *P. falciparum* 18S nested PCR (nPCR), individuals were visited weekly for clinical examination and infection status assessment by HRP2 and pLDH based RDT (RDT, First Response®, Premier Medical Corporation Ltd., Kachigam, India) microscopy and nPCR. This repeated sampling continued for up to 6 months (a total of ∼25 sampling time points). Upon detection of an incident infection by nPCR, sampling was intensified to capture the dynamics of the infection shortly after the first appearance of parasites in the blood stream and to monitor the development of high malaria parasitaemia and/or malaria symptoms that may require treatment. Finger-prick samples were taken on a daily basis for 7 days. After this first week, sampling was performed weekly until symptoms occurred (measured temperature ≥37.5°C or reported fever in last 24 hours) or, if still without symptoms, day 35. Artemether-lumefantrine treatment (AL; Coartem; Novartis Pharma) was given when symptoms occurred or on day 35, whichever came first. Upon the first detection of infection participants were invited to the insectary for assessment of infectivity; a second feed was performed on the first day of symptoms or on day 14 following first detection of parasites (whichever came first). For individuals who remained asymptomatic throughout the follow-up, a third and final membrane feeding was performed on day 35.

### Sampling: Chronic infection cohort

During screening for the chronic infection cohort, individuals were eligible for enrolment if all other eligibility criteria were met regardless of parasite status. Monthly sampling by nPCR was performed for 3-4 times, up to the peak of the transmission season. RDT’s were performed concurrently with all sampling for nPCR; positive RDT accompanied by temperature of ≥37.5°C prompted immediate treatment with AL as for incident infections. Individuals were defined as having chronic asymptomatic infection when parasites were detected by nPCR at two consecutive sampling visits, spaced one month apart, without reported illness, accompanying fever or other apparent malaria symptoms. These individuals were invited to participate in more intensive sampling. This intensive sampling phase was identical to the follow-up described after infection detection in the incident infection cohort, starting 1-2 days after the monthly visit that confirmed eligibility.

### Mosquito feeding assays

Membrane feeding was performed as described in detail in our online protocol to determine infectivity to locally reared female Anopheles *gambiae ss* mosquitoes^53^. Briefly, heparinized blood collected in vacutainers was immediately transferred to a glass mini-feeder and offered to 50-80 mosquitoes. Mosquitoes were kept at 27-29°C in the insectary on glucose and dissected 7 days after feeding. Mosquito midguts were examined for the presence of parasite developmental stages, oocysts, by two independent microscopists.

### Parasite detection and quantification

Thick blood films were stained with Giemsa and independently read by expert research microscopists over 500 fields for quantification of gametocytes and asexual parasites. The immediate molecular detection of parasites by qualitative *18S* based nested PCR (nPCR) was done at CNRFP^26^, Ouagadougou for the detection of incident and chronic infections. DNA was extracted from 20µL of blood collected on filter paper. DNA extraction was undertaken using Qiagen QIAamp blood extraction kits as per the manufacturer’s instructions (Cat No./ID: 51306). Confirmatory 18S based quantitative real-time polymerase chain reaction (qPCR) was performed at Radboudumc the Netherlands; parasite DNA was extracted from whole blood, with a detection limit of ∼0.01 parasites/µL of blood (equating to 1 parasite in each 100µL sample of extracted blood)^54^.

For subsequent analysis, total nucleic acids were extracted from EDTA blood stored in RNA protect cell reagent (Qiagen, Hilden, Germany) from 100µL blood samples stored at CNRFP at −80°C until shipment on dry ice. Extraction was done using a MagNAPure LC automated extractor (Total Nucleic Acid Isolation Kit-High Performance; Roche Applied Science, Indianapolis, IN, USA). Gametocyte quantification with qRT-PCR amplifying female (Pfs25) and male (*PfMGET, Pf3D7_1469900*) gametocyte mRNA was done as previously described, using sex-specific trendlines of cultured gametocytes^55^. Samples were declared gametocyte negative if the estimated gametocyte density was less than 0.01/µL (1 gametocyte per 100µL blood sample); estimates of male and female gametocytes were adjusted for background signal from asexual parasites ^56^. qPCR targeting *pfap2-g*^57^ and *sbp1*^58^ was performed on day of first treatment and the ratio of *pfap2-g*:*sbp1* used as an indication of the proportion of sexually committed ring-stage parasites^59^.

### Other molecular assessments

The multiplicity of infection was determined by genotyping merozoite surface protein2 (MSP2) in a nested PCR specific for MSP2 3D7 and Fc27 allelic families, followed by capillary electrophoresis (CE) to determine allelic size polymorphisms, as previously reported^60^. For MSP2-CE, samples collected on days 1, 2 and 3 of intensive follow-up were individually processed; the sum of detected clones on each of these days was combined to maximize the detectability of minority clones^61^. Human haemoglobin S (HBs) and C (HBc) were genotyped using previously published methods^62^.

### Immuno-assays

IgG antibodies against 12 antigens, 1 targeting pre-erythrocytic stages (Circumsporozoite protein [CSP]^63^) and 11 targeting the asexual blood stage (Erythrocyte binding antigen [EBA140, EBA175 and EBA181]^32^; Glutamate rich protein 2 [GLURP-R2]^64^; Merozoite surface protein 1-19 [MSP1-19]^65^, Merozoite surface protein 2 [MSP2-ch150/9 (3D7 family allele)^35^, and MSP2-DD2 (FC27 family allele)^34^]; Apical membrane antigen 1 [AMA1]^33^, and Reticulocyte binding protein homologue [RH2.2^66^, RH4.2^67^, RH5.1^68^]) were quantified at baseline for each participant using a Luminex MAGPIX© suspension bead array, as described previously ^69^. Serological analysis was conducted on serum samples collected on the first weekly scheduled visit in the incident infection cohort (serum available: n=46), or the first day of intensive daily sampling of the chronic infection cohort (serum available: n=51). Serum was assayed at a dilution of 1:200.

### Data analysis

Peak parasite densities were compared between the cohorts using Wilcoxon’s rank sum test. Changed in parasite densities over time were assessed by mixed effects linear regression on log parasite densities, with random effects for participants and a linear effect for time. Total parasite multiplication rate (PMR) was compared between different groups using mixed effects linear regression on log PMR, with random effects for participants; PMR itself is based on biological assay readouts (18s qPCR based parasite densities) so was observed to exceed 32 (the theoretical maximum PMR in one erythrocytic cycle) for a number of samples. Correlations were calculated using Spearman’s rank correlation coefficient. Associations with day 14 gametocyte densities were calculated using tobit regression on log transformed densities, with a cut off at 0.01 gametocytes per microliter. Differences in infectivity to mosquitoes was assessed binary regression with a log link. Gametocyte density was accounted for using a model previously described^24^. Repeated observations were taken into account using robust standard errors.

## Data Availability

Data will be deposited in Dryad upon publication

## Data availability

Underlying datasets are deposited in the Dryad repository upon publication.

## Acknowledgments

This work was supported by a fellowship from the European Research Council (ERC-2014-StG 639776), the Bill and Melinda Gates Foundation (INDIE OPP1173572) and the Radboud-Glasgow Collaboration Fund. JB received support from the UK MRC and the UK DFID (#MR/K012126/1) under the MRC/DFID Concordat agreement and as part of the EDCTP2 programme supported by the European Union. Recombinant proteins were kindly provided by Simon Draper (Rh5.1), James Beeson (EBA140, 175, 181, Rh2_2030 and Rh4.2), Susheel Singh (GLURP-R2), Tony Holder (MSP1-19), Mike Blackman (AMA1), Elaenor Riley (MSP2 DD2), and Kevin Marsh (MSP2 CH150/9).

## Author contributions

C.D., A.B.T and T.B. designed the study. A.B., J.B., W.S. M.M, C.D. A.B.T., and T.B. wrote the first draft of the manuscript. A.B. J.B., W.S., and T.B. analysed the data. A.B., M.W.G., A.O., I.S., I.N.O., S.S.S., K.P., S.S.A., M.O., C.W.T., D.K., Z.Z., S.B.S., A.B.T. and TB contributed to data collection in Burkina Faso. K.L., L.G. and S.S.A. developed and performed molecular assays for parasite detection and quantification. C.D., A.B.T and T.B. led the study team. All authors contributed to interpretation of the analyses and revised the draft manuscript.

## References

1 Okell, L. C., Ghani, A. C., Lyons, E. & Drakeley, C. J. Submicroscopic Infection in Plasmodium falciparum-Endemic Populations: A Systematic Review and Meta-Analysis. Journal of Infectious Diseases 200, 1509–1517, doi:10.1086/644781 (2009).

2 Ouédraogo, A. L. et al. Substantial contribution of submicroscopical Plasmodium falciparum gametocyte carriage to the infectious reservoir in an area of seasonal transmission. PLoS ONE 4, e8410 (2009).

3 Muirhead-Thomson, R. C. Factors determining the true reservoir of infection of Plasmodium falciparum and Wuchereria bacnrofti in a West African village. Transactions of the Royal Society of Tropical Medicine and Hygiene 48, 208–225 (1954).

4 Stone, W., Goncalves, B. P., Bousema, T. & Drakeley, C. Assessing the infectious reservoir of falciparum malaria: past and future. Trends Parasitol. 31, 287–296, doi:10.1016/j.pt.2015.04.004 (2015).

5 World Health Organisation. Measures of efficacy of anti-malaria interventions against malaria transmission. (WHO, Geneva, 2010).

6 Tadesse, F. G. et al. The Relative Contribution of Symptomatic and Asymptomatic Plasmodium vivax and Plasmodium falciparum Infections to the Infectious Reservoir in a Low-Endemic Setting in Ethiopia. Clin. Infect. Dis. 66, 1883–1891, doi:10.1093/cid/cix1123 (2018).

7 Ouédraogo, A. L. et al. Dynamics of the Human Infectious Reservoir for Malaria Determined by Mosquito Feeding Assays and Ultrasensitive Malaria Diagnosis in Burkina Faso. J. Infect. Dis., doi:10.1093/infdis/jiv370 (2015).

8 Hallett, R. L. et al. Chloroquine/sulphadoxine-pyrimethamine for gambian children with malaria: transmission to mosquitoes of multidrug-resistant Plasmodium falciparum. PLoS Clin Trials 1, e15, doi:10.1371/journal.pctr.0010015 (2006).

9 Targett, G. et al. Artesunate reduces but does not prevent posttreatment transmission of Plasmodium falciparum to Anopheles gambiae. Journal of Infectious Diseases 183, 1254–1259 (2001).

10 Mharakurwa, S. et al. Malaria antifolate resistance with contrasting Plasmodium falciparum dihydrofolate reductase (DHFR) polymorphisms in humans and Anopheles mosquitoes. Proc Natl Acad Sci U S A 108, 18796–18801, doi:1116162108 [pii] 10.1073/pnas.1116162108 (2011).

11 Lambrechts, L., Halbert, J., Durand, P., Gouagna, L. C. & Koella, J. C. Host genotype by parasite genotype interactions underlying the resistance of anopheline mosquitoes to Plasmodium falciparum. Malar.J. 4, 3 (2005).

12 Gouagna, L. C. et al. Genetic variation in human HBB is associated with Plasmodium falciparum transmission. Nat. Genet. 42, 328–331 (2010).

13 Bousema, T. et al. Human immune responses that reduce the transmission of Plasmodium falciparum in African populations. Int. J. Parasitol. 41, 293–300 (2011).

14 Graves, P. M., Carter, R., Burkot, T. R., Quakyi, I. A. & Kumar, N. Antibodies to Plasmodium falciparum gamete surface antigens in Papua New Guinea sera. Parasite Immunol. 10, 209–218, doi:10.1111/j.1365-3024.1988.tb00215.x (1988).

15 Graves, P. M., Doubrovsky, A., Sattabongkot, J. & Battistutta, D. Human antibody responses to epitopes on the Plasmodium falciparum gametocyte antigen PFS 48/45 and their relationship to infectivity of gametocyte carriers. Am.J.Trop.Med.Hyg. 46, 711–719 (1992).

16 Schellenberg, J. R. M. A., Smith, T., Alonso, P. L. & Hayes, R. J. What is clinical malaria? Finding case definitions for field research in highly endemic areas. Parasitology Today 10, 439–442 (1994).

17 Bousema, J. T. et al. Plasmodium falciparum gametocyte carriage in asymptomatic children in western Kenya. Malaria Journal 3, 18 (2004).

18 Dunyo, S. et al. Gametocytaemia after drug treatment of asymptomatic Plasmodium falciparum PloS Clinical Trials 1, e20 (2006).

19 Group, W. G. S. Gametocyte carriage in uncomplicated Plasmodium falciparum malaria following treatment with artemisinin combination therapy: a systematic review and meta-analysis of individual patient data. BMC Medicine 14, 79, doi:10.1186/s12916-016-0621-7 (2016).

20 Lin, J. T., Saunders, D. L. & Meshnick, S. R. The role of submicroscopic parasitemia in malaria transmission: what is the evidence? Trends Parasitol. 30, 183–190, (2014).

21 Bousema, T., Okell, L., Felger, I. & Drakeley, C. Asymptomatic malaria infections: detectability, transmissibility and public health relevance. Nat Rev Micro 12, 833–840, (2014).

22 Rono, M. K. et al. Adaptation of Plasmodium falciparum to its transmission environment. Nat Ecol Evol 2, 377–387, doi:10.1038/s41559-017-0419-9 (2018).

23 Reece, S. E., Drew, D. R. & Gardner, A. Sex ratio adjustment and kin discrimination in malaria parasites. Nature 453, 609–614, (2008).

24 Bradley, J. et al. Predicting the likelihood and intensity of mosquito infection from sex specific Plasmodium falciparum gametocyte density. eLife 7, e34463, (2018).

25 Kobayashi, T. et al. Malaria Diagnosis Across the International Centers of Excellence for Malaria Research: Platforms, Performance, and Standardization. The American Journal of Tropical Medicine and Hygiene 93, 99–109 (2015).

26 Snounou, G. et al. The importance of sensitive detection of malaria parasites in the human and insect hosts in epidemiological studies, as shown by the analysis of field samples from Guinea Bissau. Transactions of The Royal Society of Tropical Medicine and Hygiene 87, 649–653 (1993).

27 Nkhoma, S. C. et al. Close kinship within multiple-genotype malaria parasite infections. Proceedings of the Royal Society B: Biological Sciences 279, 2589–2598 (2012).

28 Wong, W. et al. Genetic relatedness analysis reveals the cotransmission of genetically related Plasmodium falciparum parasites in Thiès, Senegal. Genome Medicine 9, 5, (2017).

29 Barry, A. et al. Functional antibodies against Plasmodium falciparum sporozoites are associated with a longer time to qPCR-detected infection among schoolchildren in Burkina Faso. Wellcome Open Research 3, (2019).

30 Tran, T. M. et al. An Intensive Longitudinal Cohort Study of Malian Children and Adults Reveals No Evidence of Acquired Immunity to Plasmodium falciparum Infection. Clin. Infect. Dis. 57 (2013).

31 Rodriguez-Barraquer, I. et al. Quantification of anti-parasite and anti-disease immunity to malaria as a function of age and exposure. eLife 7, e35832 (2018).

32 Richards, J. S. et al. Association between Naturally Acquired Antibodies to Erythrocyte-Binding Antigens of Plasmodium falciparum and Protection from Malaria and High-Density Parasitemia. Clin. Infect. Dis. 51 (2010).

33 Collins, C. R. et al. Fine Mapping of an Epitope Recognized by an Invasion-inhibitory Monoclonal Antibody on the Malaria Vaccine Candidate Apical Membrane Antigen 1. J. Biol. Chem. 282, 7431–7441 (2007).

34 Taylor, R. R., Smith, D. B., Robinson, V. J., McBride, J. S. & Riley, E. M. Human antibody response to Plasmodium falciparum merozoite surface protein 2 is serogroup specific and predominantly of the immunoglobulin G3 subclass. Infection and immunity 63, 4382–4388 (1995).

35 Polley, S. D. et al. High levels of serum antibodies to merozoite surface protein 2 of Plasmodium falciparum are associated with reduced risk of clinical malaria in coastal Kenya. Vaccine 24, 4233–4246, (2006).

36 Portugal, S. et al. Treatment of Chronic Asymptomatic Plasmodium falciparum Infection Does Not Increase the Risk of Clinical Malaria Upon Reinfection. Clin. Infect. Dis. 64, 645-(2016).

37 Kakande, E. et al. Associations between red blood cell variants and malaria among children and adults from three areas of Uganda: a prospective cohort study. Malar J. 19, 21 (2020).

38 Modiano, D. et al. Haemoglobin C protects against clinical Plasmodium falciparum malaria. Nature 414, 305–308 (2001).

39 Williams, T. N. et al. Sickle cell trait and the risk of Plasmodium falciparum malaria and other childhood diseases. The Journal of infectious diseases 192, 178–186, (2005).

40 Lell, B. et al. Impact of Sickle Cell Trait and Naturally Acquired Immunity on Uncomplicated Malaria after Controlled Human Malaria Infection in Adults in Gabon. Am J Trop Med Hyg 98, 508–515 (2018).

41 Usui, M. et al. Plasmodium falciparum sexual differentiation in malaria patients is associated with host factors and GDV1-dependent genes. Nature communications 10, 2140–2140 (2019).

42 Jeffery, G. M. & Eyles, D. E. Infectivity to Mosquitoes of Plasmodium Falciparum as Related to Gametocyte Density and Duration of Infection. Am J Trop Med Hyg 4, 781–789 (1955).

43 Reuling, I. J. et al. A randomized feasibility trial comparing four antimalarial drug regimens to induce Plasmodium falciparum gametocytemia in the controlled human malaria infection model. eLife 7, e31549 (2018).

44 Brancucci, N. M. B. et al. Lysophosphatidylcholine Regulates Sexual Stage Differentiation in the Human Malaria Parasite Plasmodium falciparum. Cell 171, 1532–1544. (2017).

45 Lensen, A. et al. Plasmodium falciparum:Infectivity of Cultured, Synchronized Gametocytes to Mosquitoes. Experimental Parasitology 91, 101–103 (1999).

46 Collins, K. A. et al. A controlled human malaria infection model enabling evaluation of transmission-blocking interventions. The Journal of Clinical Investigation 128, 1551–1562 (2018).

47 Dantzler, K. W. et al. Naturally acquired immunity against immature *Plasmodium falciparum* gametocytes. Sci. Transl. Med. 11, eaav3963, (2019).

48 Gouagna, L. C. et al. Plasmodium falciparum malaria disease manifestations in humans and transmission to Anopheles gambiae: a field study in Western Kenya. Parasitology 128, 235–243 (2004).

49 Naotunne, T. S., Karunaweera, N. D., Mendis, K. N. & Carter, R. Cytokine-mediated inactivation of malarial gametocytes is dependent on the presence of white blood cells and involves reactive nitrogen intermediates. Immunology 78, 555–562 (1993).

50 Naotunne, T. S. et al. Cytokines kill malaria parasites during infection crisis: extracellular complementary factors are essential. The Journal of Experimental Medicine 173, 523–529 (1991).

51 Collins, K. A. et al. Investigating the impact of enhanced community case management and monthly screening and treatment on the transmissibility of malaria infections in Burkina Faso: study protocol for a cluster-randomised trial. BMJ Open 9, e030598, (2019).

52 Lin, J. T. et al. Microscopic Plasmodium falciparum Gametocytemia and Infectivity to Mosquitoes in Cambodia. The Journal of Infectious Diseases 213, 1491–1494, (2015).

53 Ouédraogo, A. L. et al. A protocol for membrane feeding assays to determine the infectiousness of P. falciparum naturally infected individuals to Anopheles gambiae. MWJ 4 (2013).

54 Andrews, L. et al. Quantitative real-time polymerase chain reaction for malaria diagnosis and its use in malaria vaccine clinical trials. The American journal of tropical medicine and hygiene 73, 191–198 (2005).

55 Stone, W. et al. A Molecular Assay to Quantify Male and Female Plasmodium falciparum Gametocytes: Results From 2 Randomized Controlled Trials Using Primaquine for Gametocyte Clearance. The Journal of Infectious Diseases 216, 457–467, (2017).

56 Meerstein-Kessel, L. et al. A multiplex assay for the sensitive detection and quantification of male and female Plasmodium falciparum gametocytes. Malar J. 17, 441, doi:10.1186/s12936-018-2584-y (2018).

57 Kafsack, B. F. et al. A transcriptional switch underlies commitment to sexual development in malaria parasites. Nature 507, 248–252, (2014).

58 Joice, R. et al. Inferring developmental stage composition from gene expression in human malaria. PLoS computational biology 9, e1003392, (2013).

59 Usui, M. et al. Plasmodium falciparum sexual differentiation in malaria patients is associated with host factors and GDV1-dependent genes. Nature communications 10, 2140, (2019).

60 Mueller, I. et al. Force of infection is key to understanding the epidemiology of <em>Plasmodium falciparum</em> malaria in Papua New Guinean children. Proc. Natl. Acad. Sci. U. S. A. 109, 10030–10035, (2012).

61 Koepfli, C. et al. How much remains undetected? Probability of molecular detection of human Plasmodia in the field. PloS one 6, e19010 (2011).

62 Grignard, L. et al. Bead-based assays to simultaneously detect multiple human inherited blood disorders associated with malaria. Malar J. 18, 14-14, 2648-7 (2019).

63 Kastenmüller, K. et al. Full-Length Plasmodium falciparum Circumsporozoite Protein Administered with Long-Chain Poly(I·C) or the Toll-Like Receptor 4 Agonist Glucopyranosyl Lipid Adjuvant-Stable Emulsion Elicits Potent Antibody and CD4+ T Cell Immunity and Protection in Mice. Infection and Immunity 81, 789–800, (2013).

64 Theisen, M., Vuust, J., Gottschau, A., Jepsen, S. & Høgh, B. Antigenicity and immunogenicity of recombinant glutamate-rich protein of Plasmodium falciparum expressed in Escherichia coli. Clin. Diagn. Lab. Immunol. 2, 30–34 (1995).

65 Burghaus, P. A. & Holder, A. A. Expression of the 19-kilodalton carboxy-terminal fragment of the Plasmodium falciparum merozoite surface protein-1 in Escherichia coli as a correctly folded protein. Mol. Biochem. Parasitol. 64, 165–169, (1994).

66 Triglia, T., Duraisingh, M. T., Good, R. T. & Cowman, A. F. Reticulocyte-binding protein homologue 1 is required for sialic acid-dependent invasion into human erythrocytes by Plasmodium falciparum. Mol. Microbiol. 55, 162–174 (2005).

67 Reiling, L. et al. The Plasmodium falciparum Erythrocyte Invasion Ligand Pfrh4 as a Target of Functional and Protective Human Antibodies against Malaria. PLOS ONE 7, (2012).

68 Hjerrild, K. A. et al. Production of full-length soluble Plasmodium falciparum RH5 protein vaccine using a Drosophila melanogaster Schneider 2 stable cell line system. Sci. Rep. 6, 30357 (2016).

69 Wu, L. et al. Optimisation and standardisation of a multiplex immunoassay of diverse Plasmodium falciparum antigens to assess changes in malaria transmission using sero-epidemiology. Wellcome Open Research 4, (2019).

